# An exploratory review of resiliency assessments after brain injury

**DOI:** 10.1101/2023.10.02.23296043

**Authors:** Janna Griffioen, Nicole Gingrich, Courtney Pollock, Julia Schmidt

## Abstract

**Objective:** To identify resiliency measures which have been established for use with people after acquired brain injury, using the process-based Traumatic Brain Injury Resiliency Model as the guiding conceptual framework.

**Method:** Databases CINAHL, EMBASE, Medline, and PsychINFO were searched. Using COnsensus based Standards for the selection of Health guidelines of health status Measurement INstruments (COSMIN) guidelines for reporting, articles providing data on psychometric properties for measures of resilience for people with brain injury were retrieved. Psychometric properties and clinical utility (number of items, scoring details) were summarized.

**Results:** Nine articles were retrieved, including 9 measures of resiliency.

**Conclusion:** There are established measures of resiliency in brain injury rehabilitation. Future work may explore use of these measures in a clinical context and implementation of rehabilitation goals for improving resiliency after brain injury.

## Introduction

Individuals with brain injury (e.g., stroke and traumatic brain injury) often experience long-term impacts in cognitive, physical, and psychosocial functioning [1]. People with brain injury can experience reduced participation in meaningful everyday activities and experience decreased engagement in social roles [2–5]. People with brain injury may have decreased quality of life and mental health [6,7]. For example, the presence of depression during rehabilitation has been associated with greater impairment at long term follow-up [8]. Additionally, related long-term consequences of living with brain injury can result in unemployment and quality of life [9,10]. While the experience of brain injury can initiate adversity in many facets of life, resiliency can facilitate positive processes to meet adversity after brain injury [11]. As such, resiliency may be an important construct to measure in rehabilitation.

Resilience may be lower in TBI populations compared to the general population [12]. Notably, lower resilience in TBI populations has been related to lower education and pre-injury substance use; as well as psychological distress and low quality of life experienced after a brain injury [13]. Rehabilitation that is focused on improving resiliency has shown to also improve health outcomes such as depression, anxiety, sleep, and post-traumatic stress disorder [14–16]. Resiliency also has shown impact on social outcomes like social relationships, participation, and engagement in activities after brain injury.

Resiliency is a process to negotiate cognitive, physical, and psychosocial challenges, and facilitate positive perspectives [17]. Resiliency in the context of brain injury can include the ability to appraise, adapt, and meet broad challenges associated with the consequences of brain injury [11]. The Traumatic Brain Injury Resiliency Model (TBIRM) describes processes (e.g., response to adversity, self-regulatory processes, environment) that can interact to achieve resiliency outcomes after brain injury [[11]. Resiliency has been defined in the TBIRM as, “the combined interplay among a set of affective, behavioral, and cognitive protective factors and self-regulatory processes that enable individuals to negotiate or bounce back from adversity” [11]. According to the TBIRM, resilience associates with trait or state perspectives of resilience while resiliency associates with many factors contributing to the process of resiliency [11].

Resilience and resiliency have not been consistently defined within the literature. Clinical outcomes of brain injury rehabilitation commonly focus on impairments of the injury itself (e.g., cognitive and physical challenges). While these outcomes may indicate reduction of deficits, they do not consider the influence and importance of resiliency on health and social outcomes. It is important to identify and understand measures of resiliency in brain injury in order to guide clinicians and researchers in selecting and implementing resiliency measures in clinical settings and research studies.

The purpose of this review was to identify self-report outcome measures of resiliency, validated for use with people with brain injury. The review also aimed to examine the utility of these measures in clinical rehabilitation.

## Methods

An exploratory review was conducted using the TBIRM as a conceptual framework, with a specific focus on “resiliency-related outcomes” that are subjective in nature [11]. Specifically, the TBIRM highlights three key elements when considering resiliency-related outcomes including: re-engagement of activities (e.g., participation in normal activities), adjustment of patterns (e.g., accept and adapt), and reconstruction of identity (e.g., accepting disability and creating new goals) [11].

### Sources and Search Strategy

The COnsensus based Standards for the selection of Health guidelines of health status Measurement INstruments (COSMIN) guidelines were followed as part A of the guideline, the review was conducted using four databases [18][19]. The databases were comprehensively searched since inception: CINAHL (EBSCO) 1982, EMBASE (Ovid) 1974, MEDLINE (Ovid) 1946, and PsycINFO (EBSCO) 1967 [18]. Databases were chosen as they relate to intervention by allied health groups that report on brain injury outcome measures and capture a broad interdisciplinary perspective. The last database search took place in in August 2023. Search terms were developed as indicated by COSMIN guidelines within the categories of (a) construct, (b) population, (c) clinical measure, and (d) psychometric properties of interest (see S1 Search Terms) [18]. Supporting information 1 describes the concepts used for each of the four databases. Subject headings and keyword terms that would retrieve papers on resilience and resiliency were used in combination. Combination of subject heading and keywords were combined using Boolean operators (“AND”, “OR”) were used to combine terms. Limiters for English were applied.

To obtain studies of resiliency, alternate terms were required in the database searches to broaden and increase the sensitivity of the search as the search term “resiliency” was not identified in most databases until the previous few years.

For example, EMBASE created the term resilience in 2017 from the previous term “coping behaviour”. As such, alternate terms were used to maintain a rigorous search throughout the database history. The search terms self-efficacy, acceptance, and self-confidence were identified through identifying terms used in the TBIRM, previous search terms used in the databases, librarian consultation and clinical judgement. Table 1 outlines the keyword definitions of the search terms. The search included terms for resiliency (e.g., resiliency, resilience, self-efficacy, acceptance, and self-confidence), population (e.g., acquired brain injury), clinical measure (e.g., patient reported outcome measure), and psychometric property (e.g., validity and reliability) (see S1 Search Terms).

**Table 1.** Definitions of Key Terms.

| Key Term | Definition |
| --- | --- |
| **Resiliency** | “The combined interplay among a set of affective, behavioral, and cognitive protective factors and self-regulatory processes that enable individuals to negotiate or bounce back from adversity” [p. 2709]. |
| **Resilience** | “A trait or personal quality which allows individuals to rise above or do well in spite of adversity” [11]p. 2709]. |
| **Self-confidence** | An affective component of resiliency; one’s sense of assuredness in one’s own self and abilities [[11,20]. |
| **Self-efficacy** | A behavioral component of resiliency; one’s belief in their capacity to manage a situation and achieve their desired outcome [[11,21,22]. |
| **Acceptance** | A cognitive component of resiliency; patients' acknowledgement that a problem is not likely to disappear, and it is better to adjust goals to accommodate available resources and constraints [[11,23]. |

A supplementary search was run with the title of each clinical measure, and the psychometric terms (validity and utility). The secondary search aimed to retrieve additional psychometric information specific to each clinical measure.

### Inclusion and Exclusion Criteria

Article inclusion criteria were as follows: (a) the clinical measure assessed resiliency according to the TBIRM in adults with acquired brain injuries; (b) the article included evaluation of at least one psychometric property of the given measure, even if the measure was not the focus; (c) the measure was a subjective self-report measure; (d) articles were in English, peer-reviewed full-texts. Measures that were used on acquired brain injury population (e.g., TBI stroke) were included given similar experiences with disability after injury [24].

Article exclusion criteria were as follows: (a) the measure was only validated for use with children or adolescents under the age of 18 given the different experiences of disability in a younger population [25,26]; (b) the measure was not subjective in nature, and exclusively utilized objective, proxy, or informant report information; (c) the measure does not assess resiliency in accordance to the resiliency-related outcomes in the TBIRM. If the article included collections of clinical measures, then the article was excluded. Collections of measures were further screened for any clinical measures that were missed in the search.

### Data Extraction and Analysis

The results were merged into Covidence, a systematic review management platform. Articles were screened by two independent reviewers, based on inclusion and exclusion criteria. Reviewers met to resolve conflicts after title and abstract screening, then again after full-text screening was completed (Figure).

#### **Figure.** Study Selection PRISMA

Data on each measure were extracted by two independent reviewers including population validated for use, time post-injury (i.e., community dwelling or during rehabilitation), clinical utility (i.e., number of items, size of Likert Scale, availability of clinical measure), and content of questions. Measures were evaluated for construct validity and reliability.

To understand how each measure aligns with the TBIRM, two authors independently reviewed the purpose and individual items of each measure. The measure’s purpose and items were aligned to one of the three resiliency-related outcomes of the TBIRM: re-engagement of activities, adjustment of patterns, and reconstruction of identity. In the case that some items within each measure also aligned to another resiliency-related outcomes, albeit to a lesser degree, these measures were categorized as having a secondary alignment to another resiliency outcome.

## Results

The search yielded 417 articles, leaving 305 after 112 duplicates were removed (see Figure 1). After title and abstract screening, 65 articles remained for full-text review. There were 33 articles excluded based on the inclusion criterion, with a remaining 9 articles included in this review. These articles collectively retrieved 9 clinical measures of resiliency (see Figure, for reasons of exclusion). One clinical measure, the Connor-Davidson Resilience Scale, was found from screening collections of measures such as RAND and Review of Positive Psychology Outcome Measures, but not found in the database search [1,27,28].

**Figure 1.**
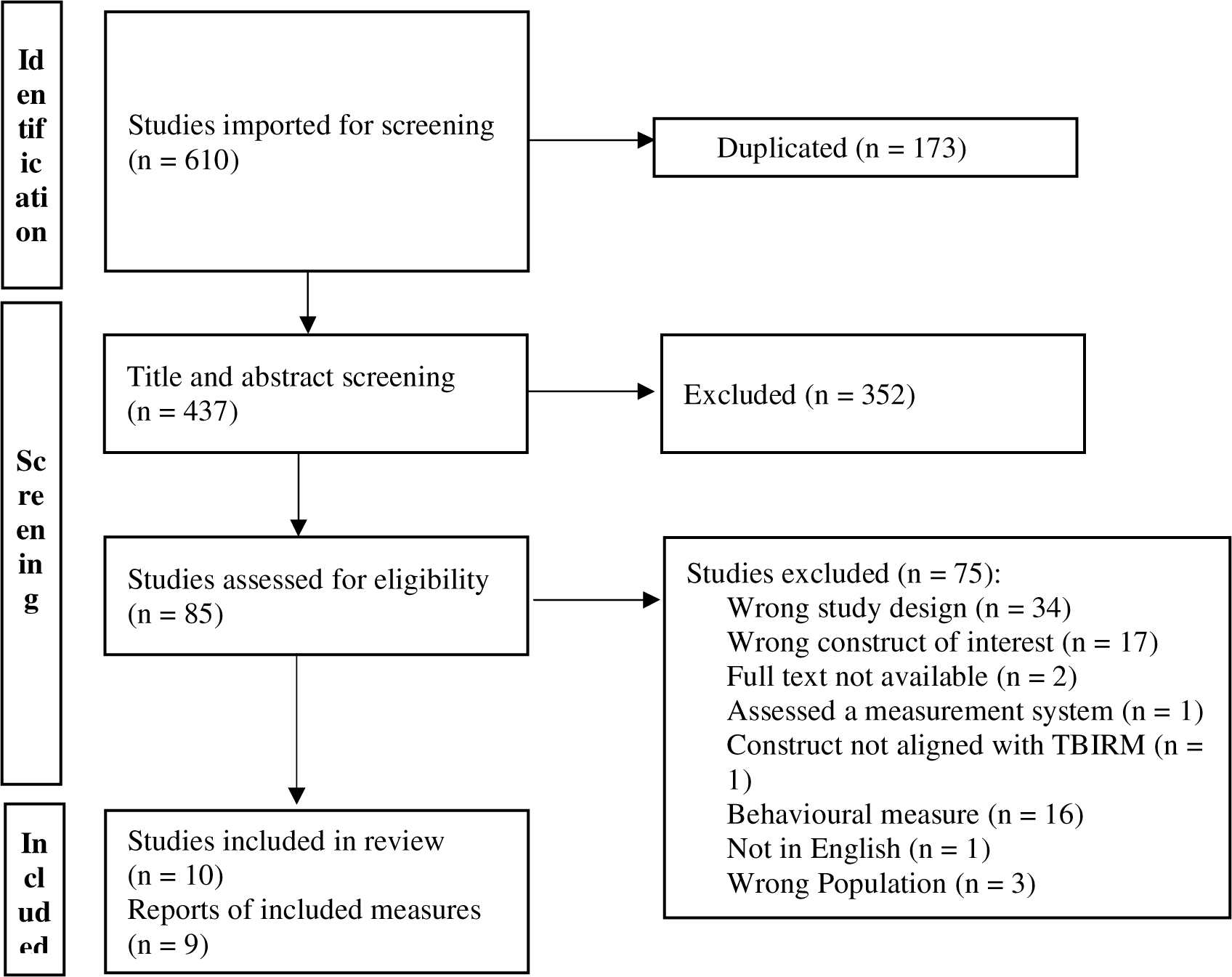
Study selection PRISMA

The measures retrieved differed in the specific clinical population which the measure has been validated for use, the format of the measure, and how the measure relates to resiliency (see Table 2). All measures incorporate a similar format of providing specific statements for participants to rate their identification with or level of agreement or disagreement with the statement. The measures are reasonable in length for ease of use clinically, in a range from 10-35 statements.

**Table 2.**
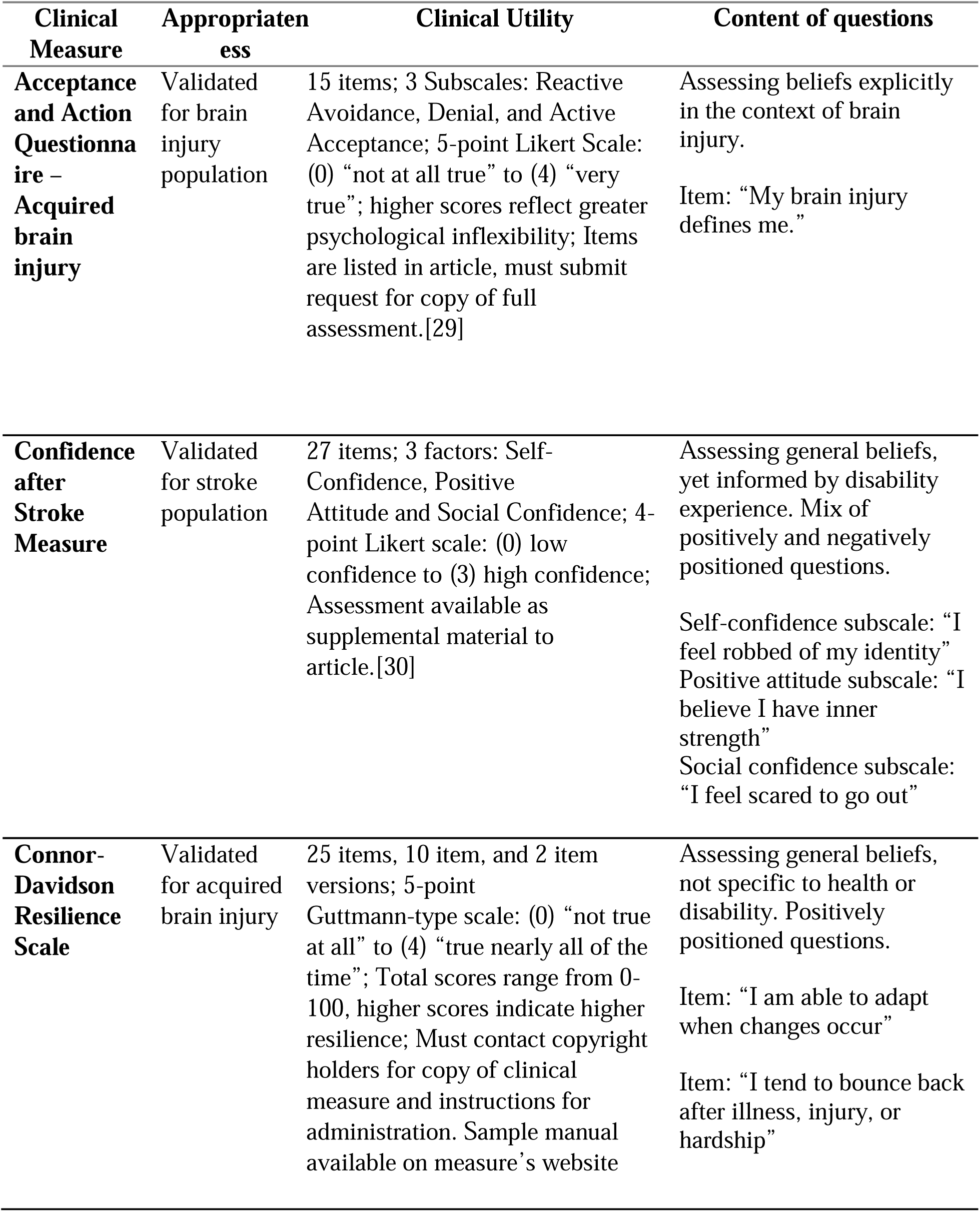
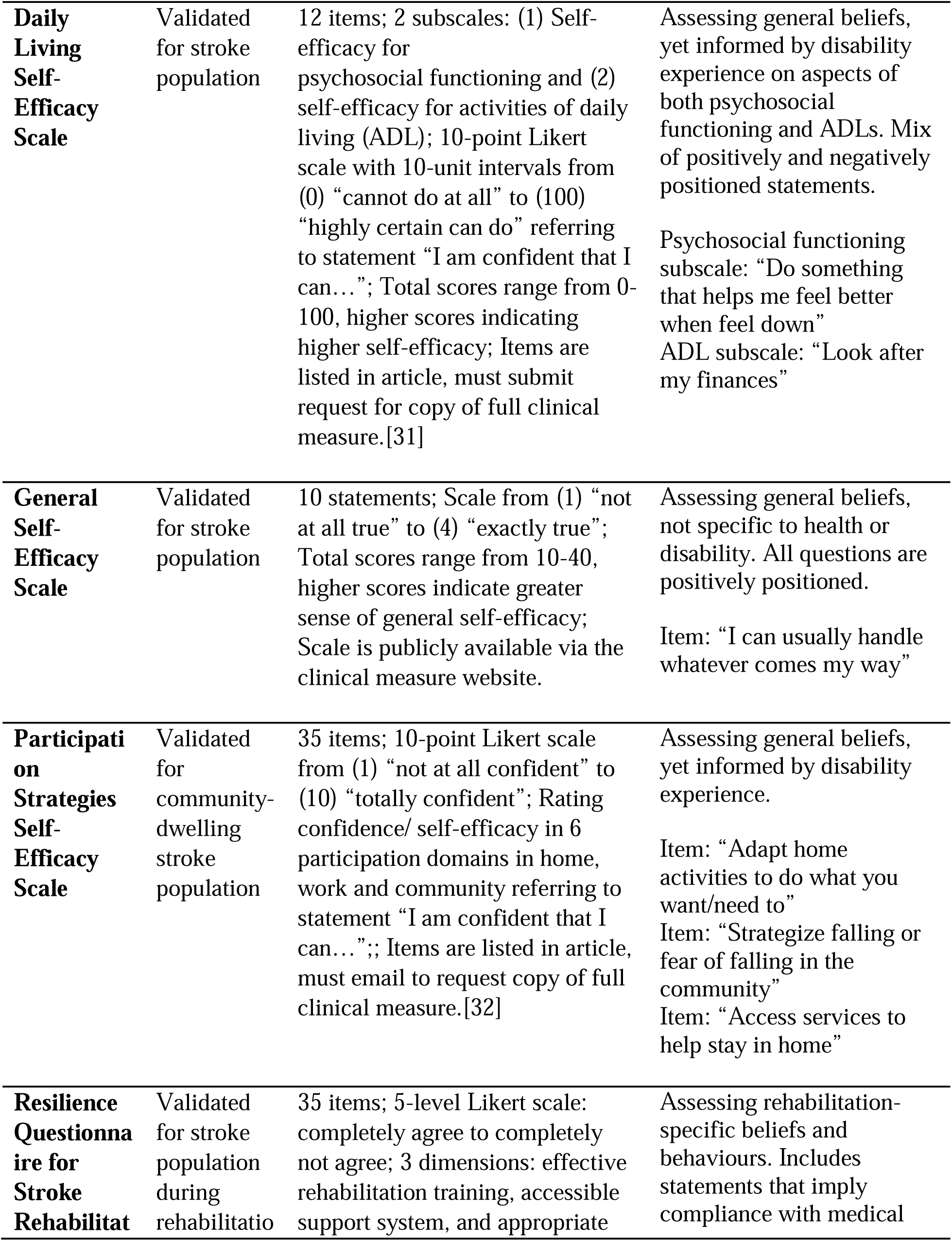
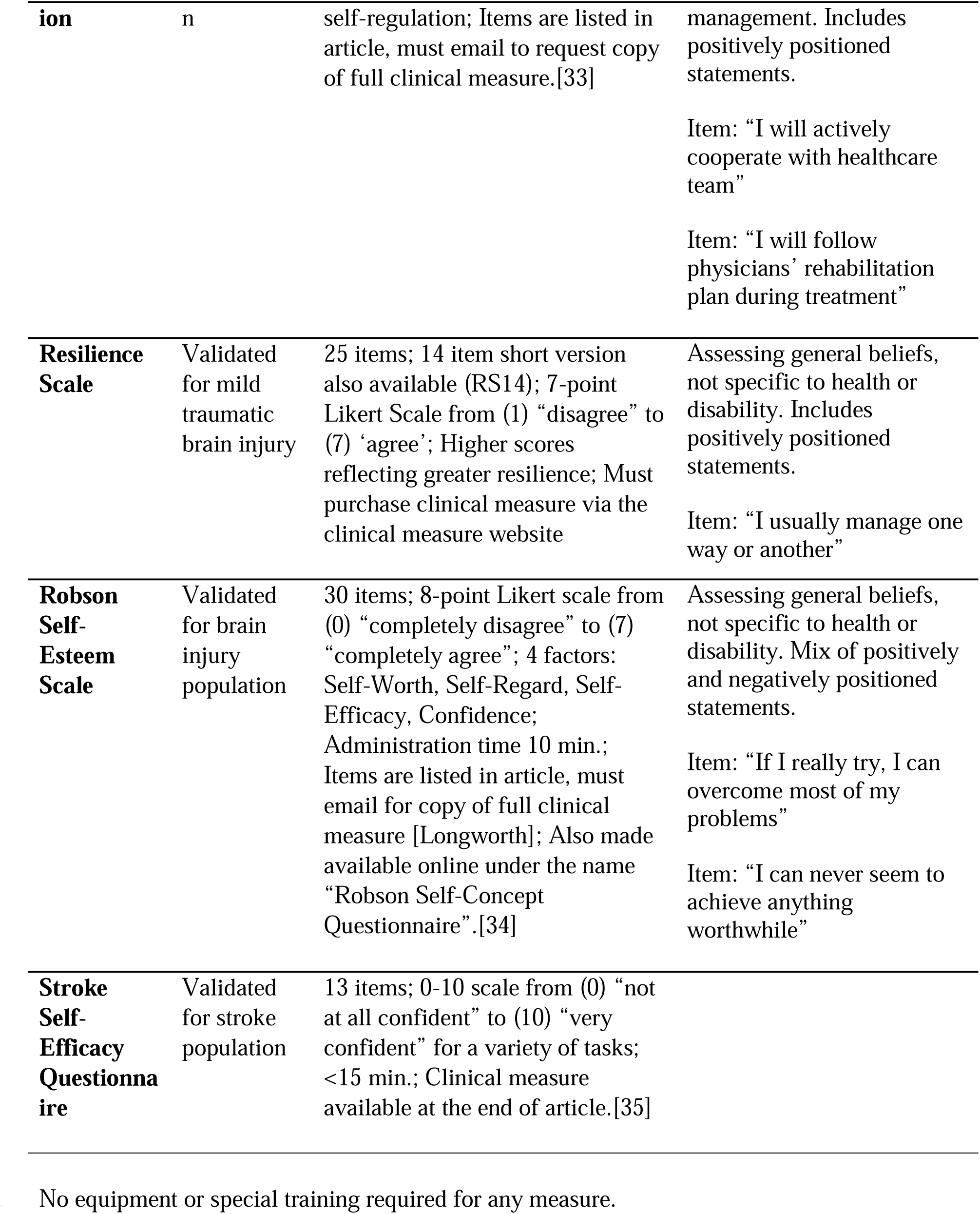
Self-Report Resiliency 171 Measures Summary Table.

### Clinical Measure

The psychometric properties of each clinical measure vary. Clinical measures in this review report psychometric properties to guide clinicians and researchers to choose appropriate measures based on the brain injury sub-population. Supporting Information 2 Psychometrics provides a summary description of the psychometric properties of each clinical outcome measure; this does not constitute an independent evaluation of psychometric properties.

### TBIRM Resiliency-Related Outcomes

Measures differed in how they related to resiliency in accordance with resiliency-related outcomes of the TBIRM (see Table 3). After analyzing the purpose and individual items of each measure, we identified four measures that considered the resiliency-related outcome of Reconstructing Identity, three measures that aligned with the outcome of Re-engaging in Activities, and two measures that assessed Adjusting Participation Patterns or Preferences. Below is a description of how each measure identified in this review aligns with the resiliency-related outcomes from the TBIRM.

**Table 3.**
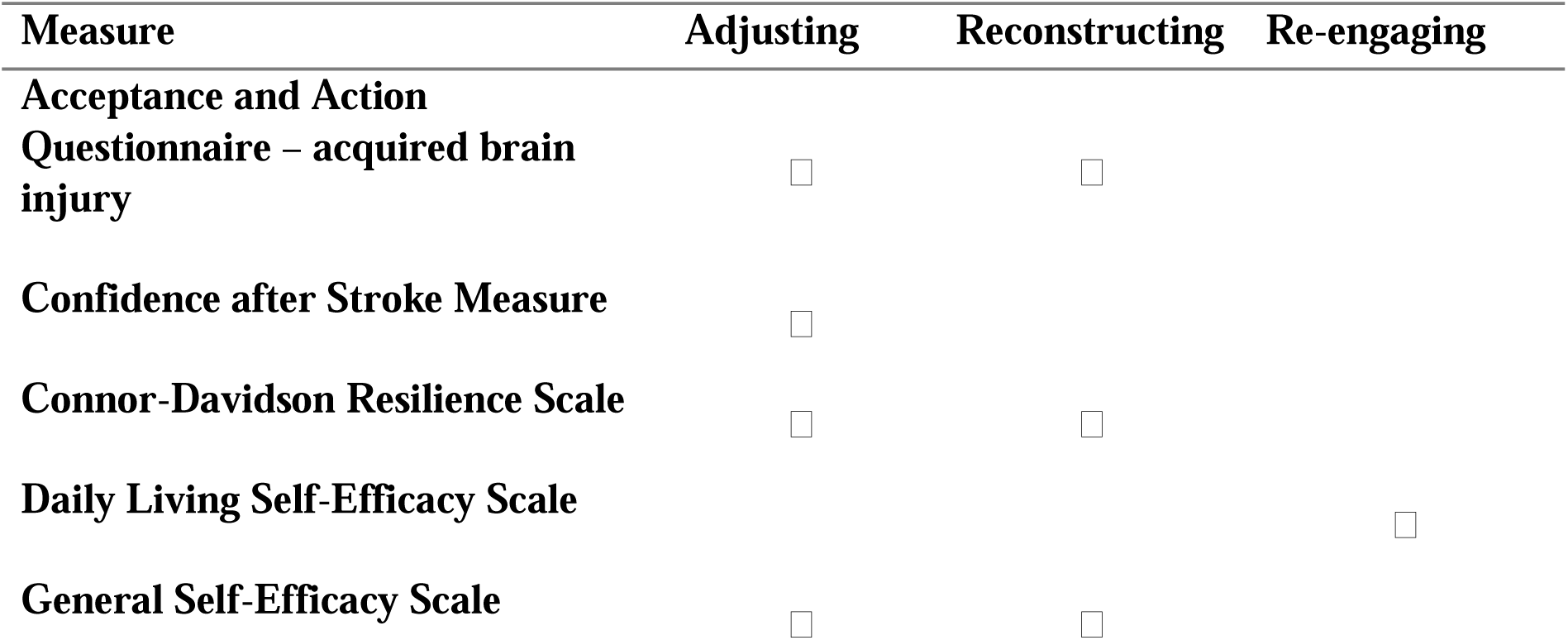
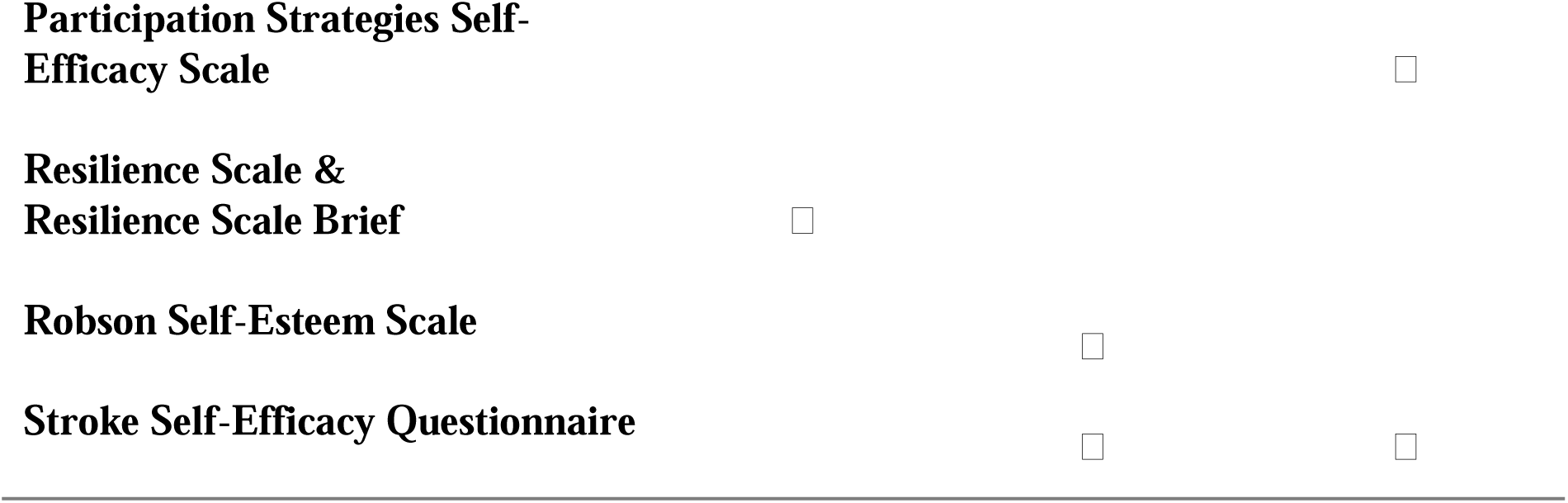
Checklist of Constructs Assessed by Measures.

### Reconstructing Identity

As per the TBIRM, reconstructing identity is understood as the ability to recognize the large change TBI enacts upon identity and the act of reconstructing identity by performance, acceptance, and identification of goals [11]. Three measures assessed the construct of Reconstructing Identity and are described below.

The General Self-Efficacy Scale is a component of the Emotional Health section of the National Institute of Health Toolbox and was validated for traumatic brain injury and stroke populations where participants were at least one-year post-injury [36]. The Swedish version has also been validated for the stroke population with good to excellent test-retest reliability and internal consistency [37]. The measure evaluates the reconstruction of identity by assessing perceived ability using questions such as “It is easy for me to stick to my aims and accomplish my goals”.

The Resilience Scale was developed for individuals 1, 6, and 12 months after mTBI, and may be limited in its applicability to community-dwelling populations with other brain injury diagnoses such as stroke to TBI [38]. The Resilience Scale evaluates the construct of reconstructing identity by using positively positioned statements that assess the individual’s identification of priorities using language that probes identity, such as “I feel that I can handle many things at a time”.

Lastly, the Robson Self-Esteem Scale, [39] assesses the process of identity reconstruction through factors such as self-deprecation, self-respect, attractiveness, and self-respect/confidence [40]. This measure has been validated for use with adults aged 17-56 who have complex and continued difficulties at least 9 months post-brain injury [39]. The measure aligned with the construct of reconstructing identity by assessing general beliefs in predominantly negatively positioned questions such as, “I can never seem to achieve anything worthwhile”.

Measures that had a secondary alignment to this outcome included the Confidence After Stroke Measure (described in full below).

### Re-engaging in Activities

The TBIRM defines re-engagement as resuming valued activities at a level equivalent to pre-injury functioning [11]. Outcome measures identified within the construct of Re-engaging Activities include the Acceptance and Action Questionnaire for Acquired Brain Injury, the Daily Self-efficacy Scale, Participation Strategies Self-Efficacy Scale, and the Stroke Self-Efficacy Scale, described below.

The Acceptance and Action Questionnaire–Acquired Brain Injury measure included 3 subscales, one of which is “Active Acceptance” [41]. In line with the definition from the TBIRM, acceptance is seen as an active process rather than a passive resignation. The measure identifies active acceptance through positive mood and relationship. The AAQ-ABI fits within the Re-engaging in Activities construct as the measure identifies active acceptance through statements such as “Even with my brain injury, I can do good work” identifying belief in ability to engage in activity with a brain injury.

The Daily Living Self-Efficacy Scale assesses self-efficacy to return to daily life with two subscales: self-efficacy for psychosocial functioning and for activities of daily living [31]. The scale has been validated with individuals several years after stroke, and its authors suggest its utility for assessing readiness for community living [31]. The scale has been validated through good to excellent test-retest reliability, convergent validity, and discriminant validity [31]. The scale includes questions like “I am confident that I can: attend a social gathering with friends” and “I am confident that I can: not allow feelings of discouragement to stop me from doing the things I want to do” which identify the process of Re-engaging in Activity as an outcome of resiliency.

Participation Strategies Self-Efficacy Scale (PSSES) assesses self-efficacy for community-level participation, with subscales for home, work, social and community management and participation [32]. The measure was developed for those with mild to moderate stroke and has limited utility in sub-acute or inpatient rehabilitation settings due to its evaluation of self-efficacy for community level participation [32]. Participation levels are measured using a subjective assessment of ability to re-engage in activity, for example “[You can] identify what you need in order to go back to work/volunteer”.

The Stroke Self-Efficacy Questionnaire, looks at self-efficacy using “self-management” and “activities” subscales and was validated with individuals 4–24 weeks post-stroke, from acute stroke units and living in the community [35,42]. The recommended utility is to assess confidence during stroke recovery and influence the approach taken by clinicians during rehabilitation [42]. The measure uses questions assessing the Re-engagement of Activity that subjectively assess capability of participating in activities of daily living, such as “Cope with the frustration of not being able to do some things because of your stroke”.

The Confidence After Stroke Measure also had a secondary alignment with the Re-engagement of Activity outcome.

### Adjusting Participation Patterns and Preferences

The TBIRM describes Adjusting Participation Patterns and Preferences as both objective adjustment and subjective adjustment [11]. Objective adjustment could be participation in new goals, while subjective adjustment could be changing perception of participation after brain injury. Outcome measures identified in this construct include the Confidence After Stroke Measure, and the Connor Davidson Resilience Scale.

The Confidence After Stroke Measure was created to guide treatment, support rehabilitation after stroke, and determine if lack of confidence is a potential barrier to recovery [34]. The measure assesses participation patterns through subjective questions “I feel less capable” and objective questions “I am confident enough to leave the house”.

The Connor Davidson Resilience Scale also fit in with Adjusting Participation Patterns and Preferences. The Connor-Davidson Resilience Scale was validated for traumatic brain injury and stroke [43] as part of the National Institute of Health Toolbox [44]. The scale has been used widely with the traumatic brain injury population [12,45–47]. The Connor-Davidson Resilience Scale is unspecific in terms of chronicity post-brain injury. The measure assesses both subjective and objective assessments of participation using positively positioned questions. The measure uses subjective phrases “I think of myself as a strong person when dealing with life’s challenges and difficulties” and objective phrases “I can deal with whatever comes my way”.

Three measures had a secondary alignment with this outcome: Acceptance and Action Questionnaire for Acquired Brain Injury, General Self-efficacy Scale, and Resilience Scale (all discussed above).

## Discussion

The present review included nine clinical measures in accordance with the TBIRM outcome measures of resiliency constructs including Reconstructing Identity, Re-engaging in Activities, and Adjusting Participation Patterns and Preferences. Of the nine clinical measures of resiliency available for use with brain injury populations, no singular clinical measure captures all aspects of resiliency-related outcomes as described by the TBIRM. This may reflect the complexity of resiliency as an outcome measure in rehabilitation as well as the need for further establishment of resiliency as important outcomes after brain injury. Further research can explore the processes of developing resiliency after brain injury as well as explore the implementation of resiliency measures in clinical practice.

This review highlights resilience as a relatively new term in library databases. As such, our search required use of terms that were related to resilience. For example, CINAHL refers to ‘hardiness’ as an alternate term to resiliency, a term that is not aligned with the TBIRM model of resiliency after TBI. Additionally, EMBASE recently created the subject heading of resilience in 2017 after previously using the term “coping behaviour”. Notably, outcome measures identified in the present review were not validated until 2009. The shift the focus from a state or trait approach (i.e., resilience) to a process perspective (i.e., resiliency) can greatly impact the field’s ability to assess resiliency from a holistic viewpoint. Theories of resiliency and models such as the TBIRM may assist with future clinical intervention and development of assessments methods.

There was variability in the applicability of the measures to different adult brain injury populations. For example, some measures were only validated for stroke populations (i.e., Confidence After Stroke Measure), others were validated for different severities and chronicity across brain injury populations (i.e., Robson Self-Esteem Scale) [39,48]. Future research could consider further development of psychometrics for each population in brain injury.

When considering clinical relevance, some measure identified hold more clinical utility than others. Clinicians should consider aspects of the measure such as population for which the measure has been validated and the stage of the person’s recovery when implementing these measures into clinical practice. While some important factors and experiences overlap among subpopulations within brain injury (e.g., stroke versus traumatic brain injury), the clinical measures included in this review have been validated for specific use. Clinicians should consider the accessibility of measures to implement in clinical practice (e.g., cost of the measure) and training required (e.g., length of training, cost of training).

Many clinical measures examine task-level domains (e.g., physical, cognitive, and emotional outcomes) important for discharge to the community and community reintegration [36,49]. Such measures may not include important patient-oriented constructs, and how resiliency influences and shapes quality of life over time [50]. The inclusion of patient-oriented constructs are important, as qualitative findings suggest that people with brain injury have alternate outcome priorities for rehabilitation after brain injury, including regaining their sense of self, improving self-efficacy, and regaining confidence in their ability to enjoy a meaningful life [35,39,42,51]. This gap in assessing patient-orientated outcomes may represent an important omission to address more explicitly during rehabilitation and community integration. Further development of patient-oriented resiliency measures could translate into clinical practice and may support clinicians and researchers to better understand the needs of people with brain injury in rehabilitation and community care [52].

The current review presents the clinical measures that have been validated in brain injury to date. The clinical measures identified in this review do not provide an exhaustive list of all resiliency measures, as the goal of this review was to consider only those validated for brain injury. The perspective of each measure focuses on the state/trait view of resilience, as opposed to the assessment of the process of resiliency. It is possible that resilience measures for other population groups (e.g., paediatrics) may be considered for further validation studies for individuals with brain injury.

## Limitations

This review had three main limitations. First, due to the small number of measures identified and associated articles, measures could not be divided into type of injury (e.g., TBI versus stroke) and time of use post injury (e.g., acute versus chronic stage of injury). However, this review provides an overview of all measures of resiliency for a brain injury population, which may have broader generalizability. However, understanding the assessments in this review may provide critical knowledge for the development of an assessment of the process of resiliency. Finally, only studies in English were included in our review which may have limited the scope of the findings, as resiliency could vary across languages and non-English speaking regions.

## Conclusion

Resiliency is an important construct to measure in brain injury rehabilitation, with many promising measures to use in this area. Further research is needed to explore how implementation of these measures in clinical practice could contribute to the development of patient goals and treatment plans directed towards the important global construct of resiliency.

## Supporting information

Supplemental 2 Psychometrics

Supplemental 1 Search Terms

Supplemental 3 PRISMA Abstract Checklist

Supplemental 4 PRISMA Checklist

## Data Availability

All relevant data are within the manuscript and Supporting Information files.

## Supporting Information Captions

S1 Appendix. Search Terms.

S2. Appendix. Psychometrics

