## Supplemental 2 Psychometrics for "An exploratory review of resiliency assessments after brain injury"

**S2. Psychometric Properties of Measures**

| Measure | Author, year | Psychometric Properties | | |
| --- | --- | --- | --- | --- |
|  |  | Participants tested | Psychometric property | Results |
| Acceptance and Action Questionnaire – Acquired Brain Injury | Kortte et al., 2009 | **n=139**  Spinal cord dysfunction secondary to spinal cord injury, Guillain-Barre, or multiple sclerosis (spinal cord): n=82;  Ischemic or hemorrhagic  strokes (stroke): n=23;  Amputations: n=16;  Hip or knee replacements (orthopedic): n=18  Age (years): Mean=54.90, SD=18.72    Male: 60.4%  Female: 39.6% | Internal consistency | a=0.70 (acceptable) |
|  |  |  | Construct validity | Facilitation of Action Factor: factor loadings = 0.16 to 0.80  Evaluation of Affect Factor: factor loadings = -0.44 to 0.69 |
|  |  |  | Predictive validity | Life satisfaction (during participation) ß=-0.45, p<0.000  Life satisfaction (3mo f/u) ß=-0.40, p<0.001  Level of handicap ß=-0.20, p<0.014  Social integration ß=-0.23, p<0.012  Rehabilitation engagement (during participation) ß=-0.13, ns  Rehabilitation engagement (3mo f/u) ß=-0.07, ns |
|  | Whiting, Diane L; Deane, Frank P; Ciarrochi, Joseph; McLeod, Hamish J; Simpson, Grahame K, 2015 | **n=150**  Severe traumatic injury: n=117; Brain tumour: n=11; Hypoxic injury: n=9; Cerebrovascular accident: n=13  Age (years): Mean=38.1, SD=13.7  Male: 77.3%  Female: 22.7% | Internal consistency | Factor 1 (Reactive Avoidance): a=0.89 (good)  Factor 2 (Denial): a=0.38 (unacceptable)  Factor 3 (Active Acceptance): a=0.46 (unacceptable) |
|  |  |  | Test-retest reliability | Factor 1 (Reactive Avoidance): ICC=0.92 (95% CI 0.86 to 0.95) (high degree of reliability)  Factor 2 (Denial): ICC=0.75 (95% CI 0.60 to 0.85) (not reliable)  Factor 3 (Active Acceptance): ICC=0.68 (95% CI 0.49 to 0.80) (not reliable) |
|  |  |  | Construct validity | Scores on Factor 1: good  Scores on Factors 2 and 3: weak |
| Confidence after Stroke Measure | Horne, Jane C.; Lincoln, Nadina B.; Logan, Pip A., 2017 | **n=202**  Healthy elderly population: n=101; Stroke respondents: n=101  Age (years): Mean=70.1, SD=13.3  Male: 45.5%  Female: 54.5% | Internal consistency (27-item questionnaire) | Participant Groups:  Stroke participants: a=0.92 (excellent)  Healthy elderly participants: a=0.90 (excellent)  Sub-scales:  Self-confidence α=0.89 (good)  Positive attitude α=0.82 (good)  Social confidence α=0.88 (good) |
|  |  |  | Test-retest reliability | Spearman’s correlation r=0.85, p=0.001 (good temporal stability)  Wilcoxon Signed Rank Test p=0.04 |
|  |  |  | Face validity | Good |
|  |  |  | Content validity | Good |
|  |  |  | Convergent validity | Spearman’s correlation between 27-item Confidence after Stroke Measure  and Stroke Self-Efficacy Questionnaire: r=0.77, p=0.001 |
| Connor-Davidson Resilience Scale | Stoner et al., 2015 | **n=806**  Random-digit dial  general population  (non-help-seeking),  primary care  recipients, psychiatric  outpatients, GAD and  PTSD  Age (years): Mean=43.8, SD not reported  Sex not reported | Internal consistency | α=0.89 (good) |
|  |  |  | Test-retest reliability | ICC=0.87 (good) |
|  |  |  | Convergent validity | With SSS: r=0.36, p<0.0001 (high significant positive correlation)  With PSS-10: r=-0.76, p<0.001 (significant negative correlation)  With SVS: r=-0.32, p<0.0001 (high significant negative correlation))  With SDS: r=-0.62, p<0.0001 (high significant negative correlation) |
|  |  |  | Criterion validity | With Kobasa hardiness: r=0.83, p<0.0001 (significant positive correlation) |
|  |  |  | Sensitivity to change | Effect of time (F=17.36; d.f. 1, 46; p<0.0001)  Interaction between time and response category F=12.87; d.f. 2, 47; p<0.001)  Both indicate scores increased with overall clinical improvement |
| Daily Living Self-Efficacy Scale | Maujean, Annick; Davis, Penelope; Kendall, Elizabeth; Casey, Leanne; Loxton, Natalie, 2014 | **n=424**  Stroke survivors: n=259; Control group (without stroke or any brain injury): n=165  Age (years): Mean=65.3, SD=12.7  Male: 46.5%  Female: 53.5% | Internal consistency | Full sample:  Total scale a=0.95 (excellent)  Psychosocial functioning a=0.94 (excellent)  Activities of daily living a=0.91(excellent)  Stroke Group:  Total scale a=0.95 (excellent)  Psychosocial functioning a=0.93 (excellent)  Activities of daily living a=0.91 (excellent)  Non-stroke Group:  Total scale a=0.88 (good)  Psychosocial functioning a=0.90 (excellent)  Activities of daily living a=0.64 (questionable) |
|  |  |  | Test-retest reliability | ICC_agreement_ of all items = 0.78-0.98 (good-excellent temporal stability) |
|  |  |  | Convergent validity | DLSES and Patient Competency Rating Scale - participants’ ratings: n=0.74, p<0.001 (high positive correlation)  DLSES and Generalized Self-Efficacy  Scale: r=0.56, p<0.001 (moderate positive correlation)  DLSES and Patient Competency Rating Scale - carers’ ratings: r=0.59, p<0.001 (moderate positive correlation) |
|  |  |  | Discriminant validity | DLSES and TICS-M: r=0.11 (non-significant correlation)  DLSES and Barthel Index: r=0.28 (very low significant positive correlation) |
| General Self-Efficacy Scale | Carlstedt, Emma, Eva Månsson Lexell, Hélène Pessah-Rasmussen, and Susanne Iwarsson. 2015 | **n=34**  Infarction (stroke): n=33; Hemorrhage (stroke): n=1  Age (years): Mean=68.1, SD not reported    Male: 61.8%  Female: 38.2% | Internal consistency | a=0.92, 95% CI 0.86 to 0.95 (excellent) |
|  |  |  | Test-retest reliability | ICC_2,1_=0.82 (95% CI 0.67 to 0.90) |
|  |  |  | Systematic/random differences | d=-0.68 (95% CI -2.23 to 0.88) |
| Participation Strategies Self-Efficacy Scale | Lee, Danbi; Fogg, Louis; Baum, Carolyn M.; Wolf, Timothy J.; Hammel, Joy, 2018 | **n=166**  Mild to moderate stroke (NIH stroke scale <16)  Age (years): Mean=56.5, SD=10.33    Male: 50.6%  Female: 49.4% | Internal consistency | Home management: a=0.904 (excellent)  Organizing at home: a=0.861 (good)  Community management: a=0.926 (excellent)  Work management: a=0.926 (excellent)  Community service management: a=0.907 (excellent)  Communication management: a=0.884 (good)  *High Cronbach’s alpha values may suggest that some items are redundant |
| Resilience Scale | Losoi, Heidi, Noah D. Silverberg, Minna Wäljas, Senni Turunen, Eija Rosti-Otajärvi, Mika Helminen, Teemu Miikka Artturi Luoto, Juhani Julkunen, Juha Öhman, and Grant L. Iverson. 2015 | **n=113**  Group 1: mild traumatic brain injury group  n=74  CT-imaged head injury patients  Age (years): Mean=37.0, SD=11.8  Male: 61%  Female: 39%  Group 2: Trauma control group n=39  Age (years): Mean=39.7, SD=12.1  Male: 49%  Female: 51% | Internal consistency | Resilience Scale:  a=0.91 to 0.93 (excellent) for mTBI group  a=0.88 to 0.95 (good-excellent) for controls  Resilience Scale-14:  a=0.88 to 0.93 (good-excellent) for mTBI group  a=0.86 to 0.94 (good-excellent) for controls |
|  |  |  | Test-retest reliability | RS across studies: 0.67-0.84  RS and RS-14 across studies: 0.66 to 0.80 |
|  |  |  | Content validity | Strong |
|  |  |  | Concurrent validity | Strong |
| Robson Self-Esteem Scale | Longworth, Catherine; Deakins, Joseph; Rose, David; Gracey, Fergus, 2018 | **n=80**  TBI: n=54; Stroke: n=18; Encephalitis: n=3; Hypoxia: n=2; Meningitis: n=1; Other: n=2  Age (years): Mean=35.55, SD=10.83    Male: 67.5%  Female: 32.5% | Internal consistency | α=0.89 (good)  Guttmann split half reliability=0.75 (good)  Factors:  Self-worth: a=0.82 (good)  Self-regard: a=0.86 (good)  Self-efficacy: a=0.72 (acceptable)  Confidence and determinism: a=0.6 (questionable) |
|  |  |  | Construct validity | Kaiser-Meyer-Olkin (KMO) measure of sampling adequacy=0.79  Bartlett’s Test of Sphericity: p<0.001  Haitovsky test: p<0.001  Factor correlation matrix: (all not significant)  Self-worth and Self-regard = -0.36  Self-worth and Self-efficacy = -0.34  Self-worth and Confidence = 0.28  Self-regard and Self-efficacy = 0.34  Self-regard and Confidence = -0.24  Self-efficacy and Confidence = -0.23 |
|  |  |  | Factorial validity | Self-Regard predicted HADS depression, accounting for 38% of variance, R^2^=0.38, F(4, 58)=9.00, p<0.001, β=−0.38, p=0.01  Two factor model:  Self-Worth (β=−0.39, p<0.01) and Self-Efficacy (β=−0.30, p<0.05) significantly predicted HADS anxiety, accounting for 44% of the variance, R2=0.44, F(4, 58)=11.26, p<0.001 |
| Stroke Self-Efficacy Questionnaire | Partridge, Cecily; Reid, Fiona; Jones, Fiona, 2008 | **n=112**  Adults with stroke  Sex not reported  Stage I  **n=15**  Age (years): Mean age and SD not reported  Stage II  **n=40**  Age (years): Mean=68.4, SD not reported  Stage III  **n=57**  Age (years): Mean=65.0, SD=17.9 | Internal consistency | a=0.90 (excellent) |
|  |  |  | Face validity | Ceiling effect for those with high degree of independence in activities of daily living and mobility, enabled 10 items to be removed from the list  Final 13-item Stroke Self-Efficacy Questionnaire had good face validity |
|  |  |  | Criterion validity | High compared with Falls Efficacy Scale, r=0.803, p<0.001 |

*CMIN/df = minimum discrepancy per degree of freedom;

CT = computed tomography;

CVI = core values index;

ICC = inter-class correlation

HADS = Hospital Anxiety and Depression Scale

MNSQ = mean square
