## Supplemental 1 Search Terms for "An exploratory review of resiliency assessments after brain injury"

S1. Search Terms

Construct: resilienc* OR self-efficacy OR self-confidence OR acceptance

AND Population: acquired brain injury OR traumatic brain injury OR brain injur* OR stroke OR cerebral vascular accident

AND Clinical measure: patient reported outcome measure OR self-report OR

outcome assessment OR outcome measure OR measure* OR assessment OR questionnaire OR rating scale

AND Psychometric properties: reliab* OR valid* OR clinical utility OR psychometric* OR psychometric properties
